# CREATION OF SAFE DOSE ZONES USING ISODOSE MAPS IN VIDEO FLUOROSCOPIC SWALLOWING STUDIES FOR PEDIATRIC PATIENTS

**DOI:** 10.1101/2024.05.14.24307144

**Authors:** P. María Ignacia Gac, H. Sofía Moncada, R. Mathias Redenz, H. Marco Jiménez, V. Ricardo Castillo

## Abstract

**Background:** During video fluoroscopic procedures in pediatric patients, a considerable number of individuals, including occupationally exposed professionals (OEP) and patient companions, must be present, exposing themselves to radiation.

**Objective:** To determine safety areas for OEP to receive the lowest absorbed dose during the examination, as shown in isodose maps.

**Method:** Technical parameters used in the examination were obtained by reviewing exams conducted during the years 2021 and 2022. Subsequently, a procedure was conducted to simulate these data using an anthropomorphic phantom and taking measurements at different points and levels of the examination room to create a map with dose zones in three dimensions. A Geiger Müller ionization chamber was used to measure the doses.

**Results:** It was found that the points closest to the patient at pelvic level of the professional had the highest doses.

**Conclusion:** OEP who must remain close to the patient have an essential obligation to comply with the use of radiological protection elements, and the relocation of elements in the room could be considered.

## Background

Swallowing video fluoroscopy, a widely used technique in the clinical setting, provides real-time dynamic images that allow for the evaluation of swallowing function in patients(1). However, its implementation involves exposure to ionizing radiation for both patients and healthcare professionals present during the procedure(2). This concern is heightened in the case of pediatric patients, where the need for diagnostic and follow-up studies is frequent, and sensitivity to radiation is greater due to their physical and metabolic development(3)(2)(4)(5).

While swallowing video fluoroscopy is invaluable for the diagnosis and management of swallowing disorders in pediatric patients(1), it is crucial to assess and mitigate the risks associated with exposure to ionizing radiation(2) This raises important questions about optimization strategies and radiation protection measures that must be implemented to ensure the safety of both patients and medical personnel during these procedures(2). There are studies that have estimated doses for both patients and professionals involved in this examination (2,6,7), concurring that efforts should continue to keep doses as low as reasonably achievable and to use radiation protection measures.

For the study of dose distribution, isodose maps have been widely used in radiation therapy treatments to assess the doses reaching the target organ and neighboring organs(8). Tests have also been conducted to evaluate the dose environment in Computed Tomography, in order to observe which areas receive higher and lower doses(9)In 2021 an article investigated through isodose maps the doses received in a city affected by radiation in order to observe through colored zones which parts are more exposed to radiation(10). In the article titled *“Radiation Protection Elements in Interventional Rooms”*(11) the distribution of doses was analyzed using isodose maps around the patient and exposed personnel, determining areas of higher dose around the patient.

In this context, the present study aims to investigate and analyze the distribution of ionizing radiation during swallowing video fluoroscopy in pediatric patients. Following the recommendations of a systematic review suggesting strategies to minimize radiation risks(2), through the construction of isodose maps and the identification of critical exposure areas, we seek to provide a deeper understanding of radiation levels present in the clinical environment during these procedures. This knowledge will enable the development of effective strategies to minimize radiation exposure and ensure the safety of patients and healthcare professionals in the context of pediatric swallowing video fluoroscopy pediatric.

## Methodology

This project has been approved by the ethics committee of our institution.

Data from video fluoroscopic swallowing examinations conducted during the years 2021-2022 were extracted from the RIS PACS AGFA system, Belgium, version 5.8.1, to extract the following technical parameters of pediatric video fluoroscopic swallowing studies in infants up to older infants (from 28 days to 24 months of age): kilovolt (kV), frames per second, series, and quantity of images per series. Sequences with images that did not contribute to the study, as well as static images or studies with excessively long sequences due to external factors, were not considered for the study. Data from 45 patients were obtained, where the mean kV was 73, the mean number of sequences was 5, and the mean number of images per sequence was 178. Subsequently, the examination was simulated using a Kyoto Kagaku PH-50 X/CT newborn anthropomorphic phantom with the mean of the parameters.

The SIEMENS AXIOM Luminos dR equipment was used for all swallow video examinations and was also utilized to set up the experiment.

For dose estimation, a Geiger Müller ionization chamber, specifically the Raysafe 452 model by Fluke Biomedicals, was employed.

A 2D floor plan of the procedure room was created using the HomeByMe program(12), depicting the various directions and distances at which the simulations were conducted (Figure 1). The creation of isodose maps was performed using Surfer 8.2 software(13).

**Figure 1.**
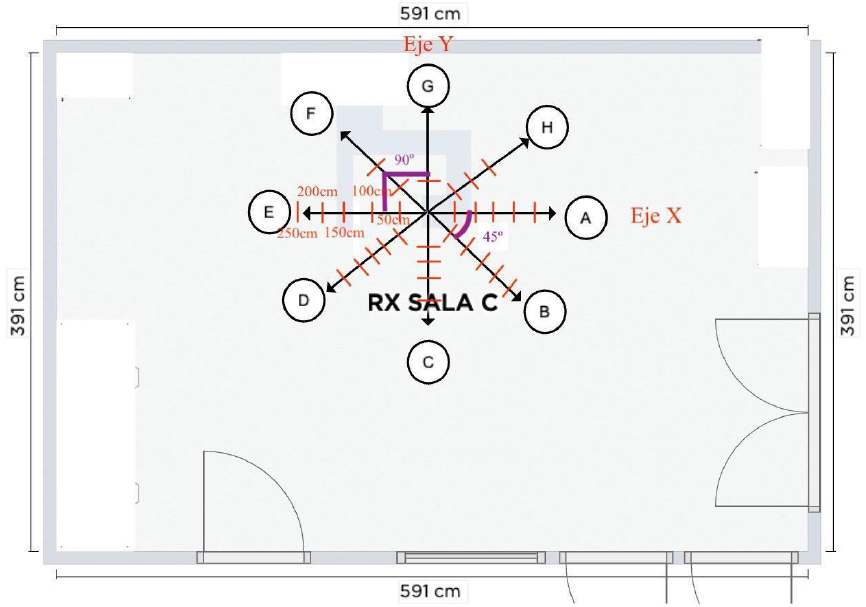
Two-dimensional layout of the procedure room *Two-dimensional layout of the procedure room and the various directions in which the simulations were conducted*.

From the same program, a 3D image was obtained to represent the three levels at which the simulations were conducted (Figure 2).

**Figure 2.**
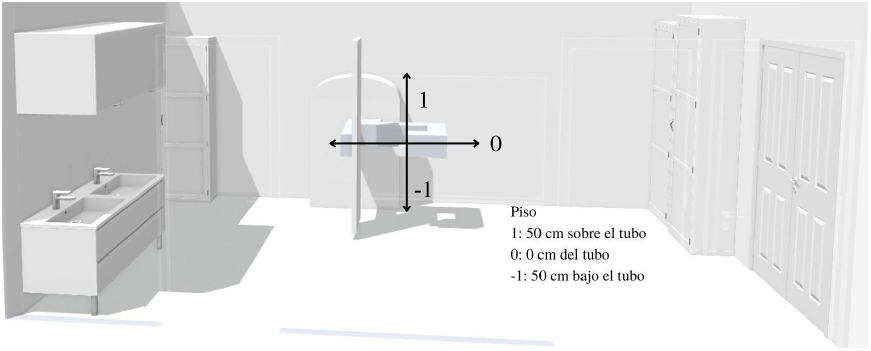
Three-dimensional layout of the procedure room *Illustration of the procedure room indicating the different levels at which the simulation was conducted*.

## Results

Once the data from the RIS-PACS system of CAS were analyzed, simulations of the examinations were conducted, yielding the following results:

**Table 1.** Cumulative dose obtained from the simulations at various points and levels.

|  | LINEA A (-1) | LINEA B (-1) | LINEA C (-1) | LINEA D (-1) | LINEA E (-1) | LINEA F (-1) | LINEA G (-1) | LINEA H (-1) |
| --- | --- | --- | --- | --- | --- | --- | --- | --- |
| 50 cm | 198 nSv* | 254 nSv | 88 nSv | 353 nSv | 281 nSv | 332 nSv | 37.7 nSv* | 178 nSv |
| 100 cm | 118 nSv | 131 nSv | 64 nSv | 329 nSv* | 240 nSv | N/A | 135 nSv | 80.8 nSv |
| 150 cm | 76.8 nSv | 88.8 nSv | 57.5 nSv | 185 nSv | 95.7 nSv | N/A | N/A | 56.9 nSv |
| 200 cm | 49 nSv | 65 nSv | 60.3 nSv | 95 nSv | 84.1 nSv | N/A | N/A | N/A |
| 250 cm | N/A | 47 nSv | N/A | 37.9 nSv | 11.4 nSv | N/A | N/A | N/A |

|  | LINEA A (0) | LINEA B (0) | LINEA C (0) | LINEA D (0) | LINEA E (0) | LINEA F (0) | LINEA G (0) | LINEA H (0) |
| --- | --- | --- | --- | --- | --- | --- | --- | --- |
| 50 cm | 9.6 nSv | 288 nSv | 518 nSv | 1220 nSv (1.22 uSv) | 6450 nSv (6.45 uSv) | 523 nSv | 226 nSv | 6.8 nSv |
| 100 cm | 11.1 nSv | 269 nSv | 318 nSv | 877 nSv | 1740 nSv (1.74 uSv) | N/A | N/A | 24.4 nSv |
| 150 cm | 10.8 nSv | 173 nSv | 220 nSv | 516 nSv | 37.1 nSv | N/A | N/A | 41.6 nSv |
| 200 cm | 10.6 nSv | 125 nSv | 147 nSv | 211 nSv | 14.3 nSv | N/A | N/A | N/A |
| 250 cm | N/A | 83.5 nSv | N/A | 110 nSv | 11.9 nSv | N/A | N/A | N/A |

|  | LINEA A (1) | LINEA B (1) | LINEA C (1) | LINEA D (1) | LINEA E (1) | LINEA F (1) | LINEA G (1) | LINEA H (1) |
| --- | --- | --- | --- | --- | --- | --- | --- | --- |
| 50 cm | 230 nSv | 212 nSv | 186 nSv | 216 nSv | 369 nSv | 198 nSv | 179 nSv | 210 nSv |
| 100 cm | 72.4 nSv | 147 nSv | 177 nSv | 285 nSv | 1330 nSv (1.33 uSv) | N/A | N/A | 142 nSv |
| 150 cm | 29.6 nSv | 91.3 nSv | 117 nSv | 180 nSv | 144 nSv | N/A | N/A | 82.5 nSv |
| 200 cm | 17 nSv | 66.3 nSv | 84 nSv | 116 nSv | 99.7 nSv | N/A | N/A | N/A |
| 250 cm | N/A | 48.8 nSv | N/A | 56 nSv | 106 nSv | N/A | N/A | N/A |

These data were input into Surfer software version 8.02, resulting in the following isodose maps:

The following map (Figure 3) was obtained from the simulation conducted at 50 cm above the floor and 50 cm below the X-ray tube. In this case, the doses did not exceed 329 nSv, achieving a more homogeneous color across the map’s extent.

**Figure 3.**
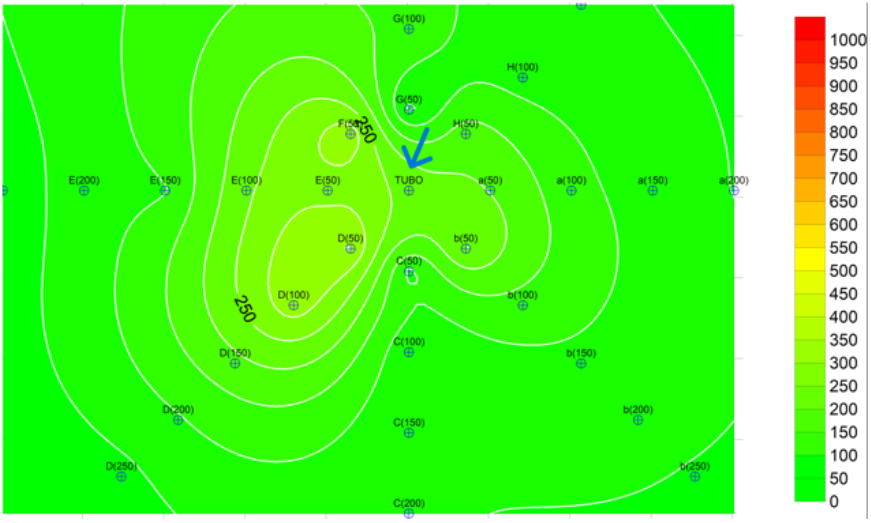
Graphic reconstruction of isodose distribution at level -1 Tube position indicated by the blue arrow. The colors on the maps indicate the amount of absorbed dose at a point in the room in nano Sieverts (nSv). Areas of lower dose are represented in green, while those of higher dose are in red.

**Figure 4.**
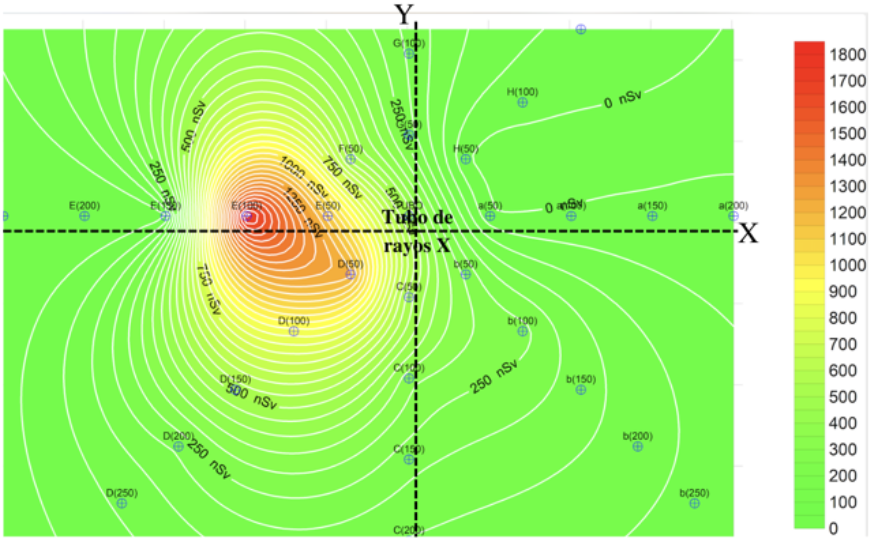
Graphic reconstruction of isodose distribution at level 0. Level 0 corresponds to points simulated at the same height as the tube, namely 100 cm above the floor. A map with the highest doses was obtained, corresponding to points near the patient of up to 6450 nSv. Points D50, D100, E50 are points within the room at common locations where speech therapists and/or radiologists position themselves to conduct the examination, and they coincide with points of highest absorbed dose.

**Figure 5.**
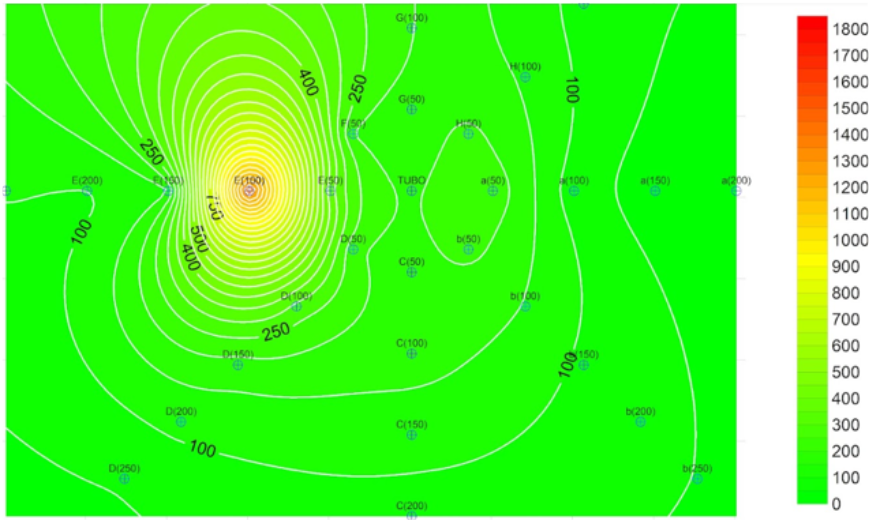
Graphic reconstruction of isodose distribution at level 1. The results from level 1, simulated at 150 cm above the floor and 50 cm above the tube, revealed high doses at the thyroid or lens level (depending on the individual’s height) in points closest to the patient. However, not as pronounced as in level 0. Additionally, the dose curve tends to increase along the Y-axis compared to the X-axis.

**Figure 6.**
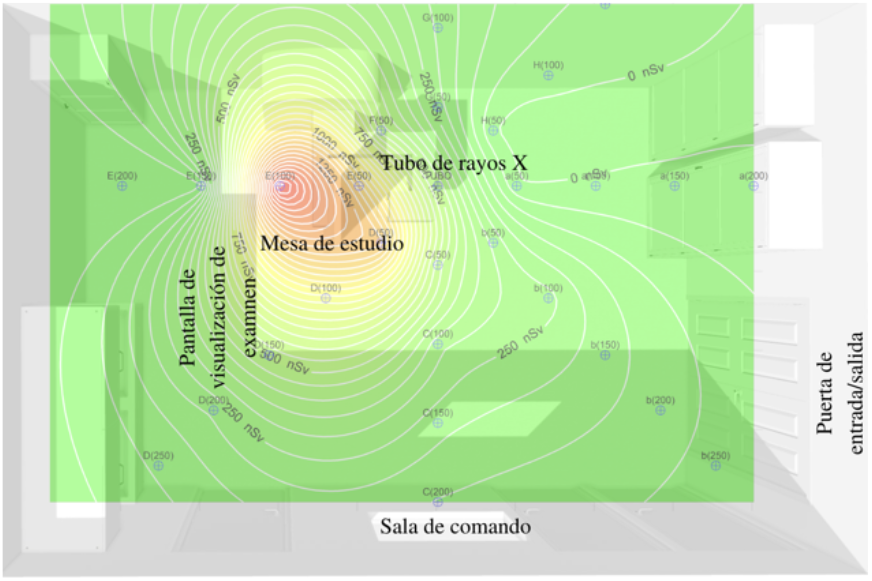
Graphic reconstruction of isodose distribution representing the layout of the room. An overlay of the isodose map from level 0 and the room layout distribution was performed to better visualize how the dose is distributed during procedures and in which directions of the room.

## Discussion

The implementation of isodose maps in the video fluoroscopy swallowing procedure room offers an invaluable tool to evaluate and understand the distribution of ionizing radiation during these clinical procedures, as evidenced by Poveda et al. 2020(11). The discussion of this strategy reveals a series of crucial findings and considerations that affect both patients and medical staff present during the video fluoroscopy swallowing procedure.

Firstly, previous studies have shown that video fluoroscopy swallowing, while essential for the diagnosis and management of swallowing disorders in pediatric patients, entails exposure to ionizing radiation(2,6,14). This concern is particularly relevant in the case of pediatric patients, whose sensitivity to radiation is greater due to their ongoing physical and metabolic development. Therefore, it is imperative to evaluate and mitigate the risks associated with radiation exposure during these procedures.

The construction of isodose maps provided a visual representation of the spatial distribution of radiation in the procedure room. These maps allow for the identification of critical exposure areas, where doses are higher, and lower-risk zones, where doses are lower, in line with the objectives of Hayano et al. 2018(10) and Kissick et al. 2024(9), who created isodose maps to determine areas of high and low doses for various purposes. When analyzing these maps, it can be observed that certain areas of the room, such as those near the patient or the X-ray tube, tend to have higher doses, while other areas, such as locations further from the point of radiation emission, show lower doses, similarly to Poveda et al. 2020(11).

The research also reveals the utility of using Surfer 8.2 software to generate isodose maps, as well as the SIEMENS AXIOM Luminos dR radiology equipment for data acquisition during video fluoroscopy procedures. These technological resources provide a robust and accurate platform for conducting detailed studies on the distribution of radiation in the clinical environment.

Furthermore, the importance of radiological protection for both patients and medical staff is highlighted(2). The results of this study underscore the need to implement effective strategies to minimize radiation exposure during video fluoroscopy swallowing. This may include optimizing the technical parameters of radiology equipment, using appropriate radiological protection elements, and implementing specific safety protocols.

In summary, the creation of isodose maps for the video fluoroscopy swallowing procedure room represents an innovative and effective approach to evaluate and manage the risks associated with ionizing radiation exposure. These maps provide valuable information that can guide clinical practice and improve the safety of patients and medical professionals during these procedures.

## Conclusions

The objective of this research project was to identify areas of lower dose within the video fluoroscopy procedure room, resulting in points A, B, C (150-200) or behind the X-ray tube being the areas of lower dose, represented in the isodose map with green color. This can recommend that in the future, when performing this examination, those present in the room have knowledge of the areas where there is a higher and lower dose of radiation, thus being able to position themselves in these areas within what is allowed during the examination. Although all participants in the examination must protect themselves with radiation protection equipment, personnel occupationally exposed who frequently conduct this examination, such as speech therapists and radiologists, must take greater precautionary measures to protect themselves from ionizing radiation, especially in areas with radiosensitive organs. The safety areas identified in this study can serve as a guide for all those present during pediatric video fluoroscopy procedures to avoid unnecessary exposure to doses. Additionally, the results obtained in this study can be used to design safety protocols for other procedures performed in this room. As for future research, the applicability of the results obtained in this study in other clinical settings and with different imaging devices could be explored. Furthermore, studies with a larger number of samples could be conducted to confirm the results obtained in this study and expand the generalizability of the results. It would also be interesting to investigate if there are differences in radiation exposure depending on the type of patient (e.g., infants and older children) and the type of procedure. This could allow the development of more specific and efficient safety protocols based on the needs and characteristics of each patient and procedure.

## Data Availability

All data produced in the present study are available upon reasonable request to the authors

## Notes

### Competing Interest Statement

The authors have declared no competing interest.

### Funding Statement

This study did not receive any funding

### Author Declarations

ethics committee Undergraduate Scientific Alemana Clinical Medicine Faculty Desarrollo University Santiago Chile approval for this work

